# Evaluating the Ruptured Arteriovenous Malformation Grading Scale (RAGS): A Reliability Study

**DOI:** 10.1101/2025.06.27.25330410

**Authors:** Jordan A. George, Sage P. Rahm, Hunter S. Boudreau, Andrew T. Hale, Travis J. Atchley, Nicholas M.B. Laskay, Philip G.R. Schmalz, Jesse Jones, Elizabeth J. Liptrap, Mark R. Harrigan, Winfield S. Fisher, Dagoberto Estevez-Ordonez

## Abstract

**Background:** Unruptured arteriovenous malformations (AVMs) carry a 1% risk of annual risk of hemorrhage; however, risk of re-hemorrhage is significantly higher after an initial rupture. Ruptured AVMs can cause significant morbidity and mortality. The Ruptured Arteriovenous Malformation Grading Scale (RAGS) was developed to better predict outcomes in patients with ruptured AVMs. However, the reliability of this scale has yet to be confirmed to support its use in clinical practice.

**Objective:** To determine the intra– and inter-rater reliability of RAGS among those most likely to use it in clinical practice.

**Methods:** A cross-sectional sample of 42 patients with ruptured AVMs was selected via retrospective review, and clinical vignettes were created. Five raters were chosen to assign a RAGS score to all 42 patients to assess the inter-rater reliability of RAGS. After two months, ten patients from the study sample were randomly selected to be re-rated to determine the intra-rater reliability of RAGS.

**Results:** The overall agreement rate was 97.2% among all raters. The inter-rater reliability was found to be substantial when measured using Cohen/Conger’s Kappa (0.73, 95% confidence interval (CI) [0.63, 0.82]), Scott/Fleiss’ Kappa (0.72, 95% CI [0.62, 0.82]), Krippendorf’s Alpha (0.73, 95% CI [.63, 95% CI [0.63, 0.82]), intraclass correlation coefficient (ICC) (0.78, 95% CI [0.68, 0.86]), and Kendall’s W (0.79, 95% CI [0.68, 0.86]) and almost perfect using Gwet’s AC (0.90, 95% CI [0.88, 0.93]). The test-retest percent agreement was between 94.7% and 98.1% among raters.

**Conclusions:** The RAGS classification system is highly reliable and has substantial to near-perfect agreement among raters with different expertise levels and specialties. This study supports the potential use of RAGS in clinical practice across different institutions.

The overall agreement rate was 97.2% among all raters. The inter-rater reliability was found to be substantial when measured using Cohen/Conger’s Kappa (0.73, 95% confidence interval (CI) [0.63, 0.82]), Scott/Fleiss’ Kappa (0.72, 95% CI [0.62, 0.82]), Krippendorf’s Alpha (0.73, 95% CI [.63, 95% CI [0.63, 0.82]), intraclass correlation coefficient (ICC) (0.78, 95% CI [0.68, 0.86]), and Kendall’s W (0.79, 95% CI [0.68, 0.86]) and almost perfect using Gwet’s AC (0.90, 95% CI [0.88, 0.93]). The test-retest percent agreement was between 94.7% and 98.1% among raters.

## Introduction

Arteriovenous malformations (AVMs) are complex vessel systems that connect arteries to veins without an intervening capillary system. Although the incidence of AVMs is relatively low, they are a significant cause of intracranial hemorrhage (ICH) in young adults. Unruptured AVMs have a 1-4% risk of hemorrhaging per year; and the risk of re-hemorrhage is about 7% after an initial rupture. ^1,2^ Management of AVMs can be complex and multidisciplinary, with options including observation, surgical resection, embolization, or stereotactic radiosurgery.

Several grading systems can assist clinicians in decision-making regarding optimal treatment. The Spetzler-Martin AVM Grading Scale (**Table 1**) estimates the risk of open surgical resection of AVMs based on size, venous drainage, and eloquence of the adjacent brain. ^3^ The Lawton-Young criteria (**Table 2**) expands upon the Spetzler-Martin scale by adding three new criteria: age, bleeding, and compactness. ^4^ These grading scales address the pre-operative risk of open surgical resection of unruptured AVMs and help guide surgical decision-making; however, no grading scale is designed to estimate outcomes after an AVM rupture. The Ruptured Arteriovenous Malformation Grading Scale (RAGS) (**Table 3**) has been implemented to predict outcomes in patients with ICH due to AVM rupture. ^5^ RAGS is a nominal scale incorporating the Hunt-Hess grading scale (**Table 4**). ^6^ The reliability of RAGS has not yet been determined. In this study, we present an evaluation of the inter-rater and intra-rater reliability of the RAGS.

**Table 1.** Spetzler-Martin Arteriovenous Malformation Grading Scale.

| Grading Factor | Criteria | Points |
| --- | --- | --- |
| Size of AVM | Small (< 3 cm) | 1 |
|  | Medium (3 – 6 cm) | 2 |
|  | Large (>6 cm) | 3 |
| Pattern of Venous Drainage | Superficial only | 0 |
|  | Deep involvement (e.g. deep veins) | 1 |
| Eloquence of Adjacent Brain | Non-eloquent area | 0 |
|  | Eloquent Area | 1 |
| Grade | Surgical Risk |  |
| I (1 Point) | Low |  |
| II (2 Points) | Moderate |  |
| III (3 Points) | Moderate-to-High |  |
| IV (4 Points) | High |  |
| V (5 Points) | Very High |  |

**Table 2.**
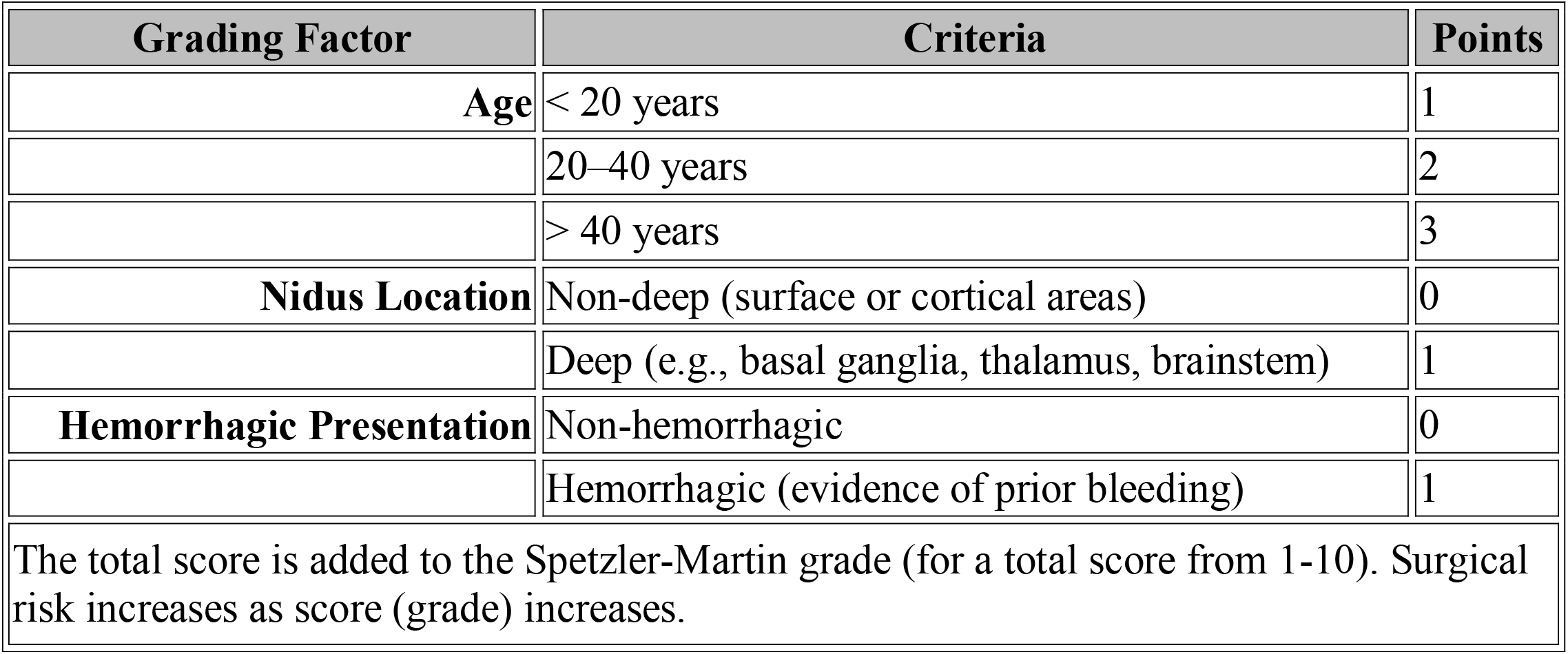
Lawton-Young Supplemental Grading System.

**Table 3.** Ruptured AVM Grading Scale (RAGS)

| Parameter | Score | Description |
| --- | --- | --- |
| **Age** | 0 | < 40 years |
|  | 1 | ≥ 40 years |
| **Hunt and Hess Grade** | 0 | Grades 1–2 |
|  | 1 | Grades 3–5 |
| **Intraventricular Hemorrhage (IVH)** | 0 | Absent |
|  | 1 | Present |
| **Deep AVM Location** | 0 | Absent (cortical or superficial location) |
|  | 1 | Present (e.g., basal ganglia, thalamus, or brainstem) |
| **Size of AVM** | 0 | < 3 cm diameter |
|  | 1 | ≥ 3 cm diameter |
| **Total RAGS Score** | **Prognosis** |  |
| **0-2** | Favorable outcome likely |  |
| **3-4** | Intermediate prognosis |  |
| **5** | Poor prognosis, high morbidity risk |  |

**Table 4.** Hunt and Hess Grading System.

| **Grade** | **Clinical Condition** | **Prognosis** |
| --- | --- | --- |
| **1** | Asymptomatic or mild headache with slight nuchal rigidity | Excellent prognosis |
| **2** | Moderate to severe headache, nuchal rigidity, and no neurological deficits (except cranial nerve palsy) | Good prognosis |
| **3** | Drowsiness, confusion, or mild focal neurological deficit | Fair prognosis |
| **4** | Stupor with moderate to severe hemiparesis, early decerebrate rigidity, and vegetative disturbances | Poor prognosis |
| **5** | Deep coma, decerebrate posturing, moribund appearance | Very poor prognosis; high mortality |

## Methods

A cross-sectional single-institutional sample of 42 patients with ruptured AVMs was selected via retrospective review based on an *a priori*-determined sample size calculation. Institutional review board (IRB) approval was obtained prior to accessing patient data. Patient consent was not required as only the patient’s imaging and electronic medical record were accessed, for which IRB approval is sufficient. Clinical vignettes consisting of the relevant patient history, physical exam findings, and imaging studies (computed tomography (CT), CT angiogram (CTA), and/or digital subtraction angiography (DSA)) were created. The patient cohort comprised the entire spectrum of possible RAGS values. Inclusion criteria were patients >18 years of age with ruptured AVMs treated at our single institution between 2000 and 2022 with necessary imaging studies to assign a RAGS value. Exclusion criteria included patients <18 years of age and those with ruptured AVMs lacking the imaging required to assign a RAGS score. Five raters (two dual-trained vascular neurosurgeons, two senior neurosurgery residents, and one neurointerventional radiologist) were selected to assign a RAGS value to all 42 patients to assess the inter-rater reliability of RAGS. After two months, ten patients from the study sample were randomly selected to be re-rated to evaluate the intra-rater reliability of RAGS.

This manuscript adheres to the Guidelines for Reporting Reliability and Agreement Studies (GRRAS) promoted by the EQUATOR Network. ^7^

### Statistical Analysis

The sample size was calculated based on an expected ICC of 0.6 (H0 p = p0) with five raters and a desired power of 0.8 with alpha set at 0.05. We chose p0 = 0.4 as the minimal acceptable level of reliability and null value to perform a one-tailed test of significance for each calculated coefficient (H0: coefficient <= 0.4000). Statistical analyses were performed using STATA/MP version 18 (StataCorp, College Station, TX) and R Statistical Software (v4.1.2; R Core Team 2021). The kappaetc community-contributed program available for use with STATA was used to calculate percent agreement, weighted Conger’s kappa, weighted Fleiss’ generalized kappa for multiple raters, Krippendorff’s alpha, and interclass correlation coefficient (ICC). ^8-11^ We assumed a 2-way random effects model with individual scoring from each rater and employed the Shrout and Fleiss ICC Model 2 (ICC2,1) to analyze absolute agreement. ^12^ Kendall’s concordance coefficient (W) was calculated using the IRR R package (v0.84.1). Using Efron’s approach, confidence intervals for Kendall’s W were obtained through bootstrap resampling with 10,000 generated samples. ^13^ Interpretation of agreement was done through standard methodology proposed for each measurement. ^9-11^ The data and code used for data analysis in this report have been made available publicly through GitHub (https://github.com/estevezdo/RAGS_Reliability_Study).

## Results

Per GRRAS reporting guidelines, the results below were based on five raters (two dual-trained vascular neurosurgeons, two senior neurosurgery residents, and one neurointerventional radiologist).

### Interrater Reliability

The overall percent agreement was 97.2% among all raters. The inter-rater reliability was found to be substantial when measured using Cohen/Conger’s Kappa (0.73, 95% confidence interval (CI) [0.63, 0.82]), Scott/Fleiss’ Kappa (0.72, 95% CI [0.62, 0.82]), Krippendorf’s Alpha (0.73, 95% CI [0.63, 95% CI [0.63, 0.82]), intraclass correlation coefficient (ICC) (0.78, 95% CI [0.68, 0.86]), and Kendall’s W (0.79, 95% CI [0.68, 0.86]) and almost perfect using Gwet’s AC (0.90, 95% CI [0.88, 0.93]). These results were all statistically significant and listed in **Table 5**.

**Table 5.** RAGS Inter-rater Reliability Estimates. TKendall’s coefficient of concordance (W) for ordinal data; aclass correlation (ICC) for ordered, Likert-type, or continuous data.

| Statistical Measure | Coefficient | 95% Confidence Interval |
| --- | --- | --- |
| Cohen/Conger's Kappa | .73 | 0.63, 0.82 |
| Scott/Fleiss' Kappa | .72 | 0.62, 0.82 |
| Krippendorff's Alpha | .73 | 0.63, 0.82 |
| ICC | .78 | 0.68, 0.86 |
| Kendall's W | .79 | 0.68, 0.86 |
| Gwet's AC | .90 | 0.88, 0.93 |

### Intrarater Reliability

The test-retest percent agreement was between 94.7% and 98.1% among raters. The intra-rater reliability was found to be substantial when measured using Cohen/Conger’s Kappa (0.68-0.87), Scott/Fleiss’ Kappa (0.68-0.87), and Krippendorf’s Alpha (0.70-0.88), and almost perfect using Gwet’s AC (0.83-0.93), ICC (0.79-0.91), and Kendall’s W (0.82-0.91). These results were all statistically significant and shown in **Table 6** and across wide range of possible RAGS values (**Figure 1**)

**Table 6.** RAGS Intra-rater Reliability Estimates. Kendall’s coefficient of concordance (W) for ordinal data; aclass correlation (ICC) for ordered, Likert-type, or continuous data.

|  | Cohen/Conger's<br>Kappa | Scott/Fleiss'<br>Kappa | Krippendorff's<br>Alpha | ICC | Kendall's<br>W | Gwet's<br>AC |
| --- | --- | --- | --- | --- | --- | --- |
| Rater 1 | .68 | .68 | .70 | .79 | .83 | .82 |
| Rater 2 | .74 | .72 | .73 | .82 | .91 | .85 |
| Rater 3 | .79 | .78 | .80 | .83 | .87 | .87 |
| Rater 4 | .87 | .87 | .88 | .91 | .90 | .93 |
| Rater 5 | .76 | .76 | .77 | .81 | .88 | .87 |

**Figure 1.**
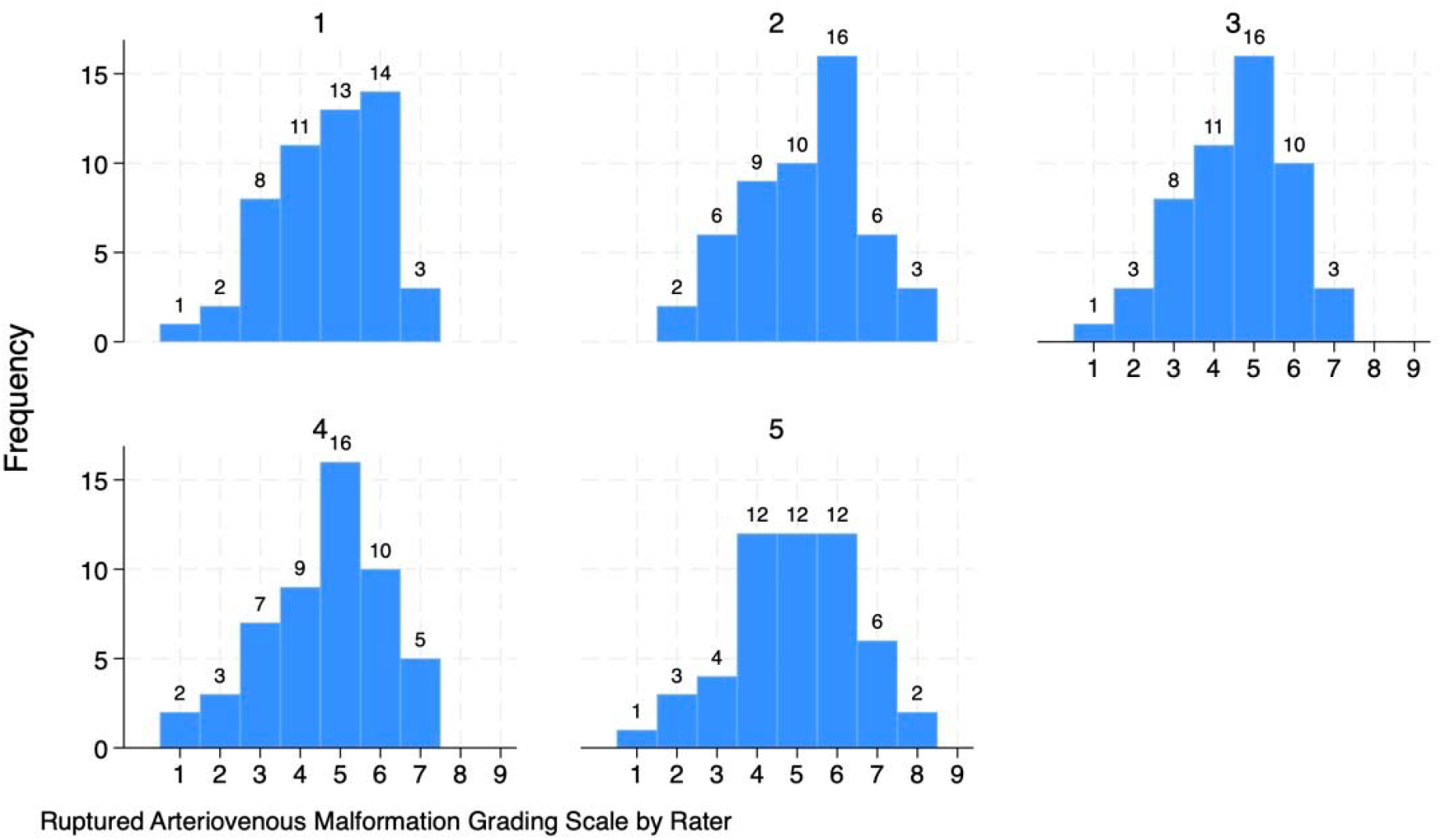
Frequency of RAGS values assigned per rater.

## Discussion

Traditionally, AVMs have been classified based on size, deep venous drainage, and eloquence to determine the risk associated with open surgical resection. ^3^ The Spetzler-Martin and Lawton-Young grading scales have shown substantial reliability and are commonly employed in clinical practice.

However, until recently, there has been no grading scale to predict outcomes in patients with ruptured AVMs accurately. The RAGS is a promising new grading scale that attempts to predict outcomes in these patients; however, to date, no studies have assessed its reliability. Here, we report substantial to near-perfect inter– and intra-rater reliability and provide data that supports its use in clinical practice and trials.

The overall percent agreement was 97.2% among all raters. Inter-rater reliability was substantial (using Cohen/Conger’s Kappa, Scott/Fleiss’ Kappa, Krippendorf’s Alpha, ICC, and Kendall’s W) to almost perfect (using Gwet’s AC).

The RAGS intra-rater reliability showed similar results. The test-retest percent agreement was between 94.7% and 98.1% among raters. Intra-reliability was substantial when measured with Cohen/Conger’s Kappa, Scott/Fleiss’ Kappa, Krippendorf’s Alpha, and ICC, and almost perfect with Gwet’s AC and Kendall’s W. These proved true over a wide range of possible RAGS values. (**Figure 1)**.

Although reliability is important, validity for RAGs is just as crucial. Silva et al. demonstrated that RAGS estimate long-term clinical outcome more accurately than other grading systems in 115 patients with ruptured AVMs (mean follow up of 4 years; AUROC > 0.80 across all follow-up periods).^5^ A multicenter retrospective study examining modified Rankin Scale (mRS) outcomes of 61 adults with ruptured AVMs and found that RAGS was a reliable scale predicting long-term outcomes in adults up to a year after rupture (0.79, 0.76, and 0.73 for dichotomized mRS at the first 6 months, between 6 and 12 months, and after 12 months of follow-up, respectively).^14^

RAGS has been found to be valid for pediatric patients as well. A retrospective study found that RAGS outperformed other clinical grading scales in predicting long-term functional outcomes assessed by mRS in 81 children with ruptured AVMs (mean follow up of 4 years; AUROC > 0.80 across all follow-up periods).^15^ These three studies support the scale’s ability to predict long-term disability and survival in both pediatric and adult patients with ruptured AVMs.

Ultimately, the utility of the RAGS classification system is reinforced by the high inter– and intra-reliability found in this study. Additionally, the scale’s simplicity makes it accessible for clinical use, even in resource-limited settings. By incorporating variables such as patient age, Hunt and Hess grade, presence of intraventricular hemorrhage, AVM size, and location, RAGS offers a practical framework to stratify prognosis and guide management decisions for patients with ruptured AVMs.

## Limitations

This study was conducted at a single institution. Other centers that may not have specialized neurovascular neurosurgeons or resident physicians assessing patients with ruptured AVMs, and therefore may have different degrees of reliability utilizing RAGS. At many academic institutions with resident physicians, “junior” residents are often involved in the evaluation and management of patients with ruptured AVMs. Although “senior” residents were involved in the rating process of this study, “junior” residents were not included. Residents with less experience may potentially lower the reliability of RAGS. Due to the retrospective study design, reviewers determined Hunt-Hess scores (HH scores) via chart review rather than physical exam. The variability of neurological exam descriptions in patient charts may have contributed to increased variation in both HH scores and consequently RAGS. Ultimately, this study evaluates the inter– and intra-rater reliability of RAGS but does not give any insight on its validity. Although there is excellent reliability, prospective studies are needed to reinforce RAGS as a useful tool to predict clinical outcomes in patients with ruptured AVMs.

## Conclusion

This study demonstrates that the RAGS classification system is a highly reliable clinical tool. To date, this is the first external reliability study of this grading system. Further studies are needed to evaluate the role of RAGS in guiding clinical management and assessing its validity in predicting long-term outcomes of patients with ruptured AVMs.

## Data Availability

All data produced in the present work are contained in the manuscript.

## Notes

Conflict of interest: None

### Competing Interest Statement

The authors have declared no competing interest.

### Funding Statement

None.

### Author Declarations

The University of Alabama at Birmingham Institutional Review Board approved this study.

## References

1. Lawton MT, Rutledge WC, Kim H, et al. Brain arteriovenous malformations. Nat Rev Dis Primers. May 28 2015;1:15008. doi:10.1038/nrdp.2015.8

2. Ondra SL, Troupp H, George ED, Schwab K. The natural history of symptomatic arteriovenous malformations of the brain: a 24-year follow-up assessment. J Neurosurg. Sep 1990;73(3):387–91. doi:10.3171/jns.1990.73.3.0387

3. Spetzler RF, Martin NA. A proposed grading system for arteriovenous malformations. J Neurosurg. Oct 1986;65(4):476–83. doi:10.3171/jns.1986.65.4.0476

4. Hafez A, Koroknay-Pál P, Oulasvirta E, et al. The Application of the Novel Grading Scale (Lawton-Young Grading System) to Predict the Outcome of Brain Arteriovenous Malformation. Neurosurgery. Feb 1 2019;84(2):529–536. doi:10.1093/neuros/nyy153

5. Silva MA, Lai PMR, D. R, Aziz-Sultan MA, Patel NJ. The Ruptured Arteriovenous Malformation Grading Scale (RAGS): An Extension of the Hunt and Hess Scale to Predict Clinical Outcome for Patients With Ruptured Brain Arteriovenous Malformations. Neurosurgery. Aug 1 2020;87(2):193–199. doi:10.1093/neuros/nyz404

6. Hunt WE, Hess RM. Surgical risk as related to time of intervention in the repair of intracranial aneurysms. J Neurosurg. Jan 1968;28(1):14–20. doi:10.3171/jns.1968.28.1.0014

7. Kottner J, Audige L, Brorson S, et al. Guidelines for Reporting Reliability and Agreement Studies (GRRAS) were proposed. Int J Nurs Stud. Jun 2011;48(6):661–71. doi:10.1016/j.ijnurstu.2011.01.016

8. Klein D. Implementing a General Framework for Assessing Interrater Agreement in Stata. The Stata Journal. 2018;18(4):871–901. doi:10.1177/1536867x1801800408

9. Fleiss J. The Design and Analysis of Clinical Experiments. 2010;doi:10.1002/9781118032923

10. Landis JR, Koch GG. The measurement of observer agreement for categorical data. Biometrics. Mar 1977;33(1):159–74.

11. Kendall M, Gibbons J. Rank Correlation Methods. Oxford University Press; 1990.

12. Koo TK, Li MY. A Guideline of Selecting and Reporting Intraclass Correlation Coefficients for Reliability Research. J Chiropr Med. Jun 2016;15(2):155–63. doi:10.1016/j.jcm.2016.02.012

13. Efron B. Better Bootstrap Confidence Intervals. J Am Stat Assoc. 1987;doi:10.1080/01621459.1987.10478410

14. Antkowiak L, Rogalska M, Stogowski P, et al. External validation of the Ruptured Arteriovenous Malformation Grading Scale (RAGS) in a multicenter adult cohort. Acta Neurochir (Wien). Apr 2023;165(4):975–981. doi:10.1007/s00701-022-05433-1

15. Garcia JH, Rutledge C, Winkler EA, et al. Validation of the Ruptured Arteriovenous Malformation Grading Scale in a pediatric cohort. J Neurosurg Pediatr. May 1 2022;29(5):575–579. doi:10.3171/2022.1.Peds21466

